# Perspectives on Cervical Cancer Prevention Strategies and a Combination Treatment for Cervical Precancer in South African Women Living with HIV and Male Partners

**DOI:** 10.1101/2025.01.07.25320108

**Authors:** Nicholas S. Teodoro, Nkosinathi Ngcobo, Milenka Jean-Baptiste, Masangu Mulongo, Cecilia Milford, Malgorzata Beksinska, Jaqueline Burgess, Sibusisiwe Luvuno, Jermina Nkoana, Nonkululeko Mayisela, Lisa Rahangdale, Carla J. Chibwesha

## Abstract

**Introduction:** Cervical cancer remains the second most common cancer among women worldwide, with 85% of cases occurring in low-and middle-income countries (LMIC). Women living with HIV (WLWH) are at a particularly high risk of developing for high-grade cervical intraepithelial neoplasia (CIN2/3) and cervical cancer, and the standard surgical treatment is far less effective in this population. Thus, research on medical therapies and combination treatment options remain a priority. In preparation for a clinical trial involving adjuvant intravaginal 5-fluorouracil (5FU) cream following surgical treatment for CIN2/3, we explored the acceptability of our proposed intervention among WLWH and male partners.

**Methods:** We conducted qualitative interviews with WLWH and male partners in Johannesburg, South Africa between April 2022 and September 2022. We invited WLWH to participate in semi-structured focus group discussions (FGDs), while male partners were invited to participate in semi-structured in-depth interviews (IDIs). The analysis utilized a rapid, deductive approach in which quotations were identified and categorized into relevant domains: factors affecting cervical cancer screening, the initiation of 5FU vaginal cream, and adherence to 5FU.

**Results:** We conducted 9 FGDs comprising 48 WLWH and 18 IDIs with male partners. The mean age of participants was 43 years, and the majority (75%) had completed secondary education. Most women (75%) had also undergone Pap smear screening and 50% had a prior abnormal Pap smear. Qualitative analysis revealed that education on HPV and cervical cancer, prior experience with the health system, and social perceptions/stigma influenced cervical cancer screening uptake and were important factors in the initiation of and adherence to 5FU. Men’s knowledge about cervical cancer was extremely limited. Overall, participants’ perceptions of the proposed trial intervention were positive, with most participants expressing confidence that women would be able to use and adhere to the intravaginal 5FU cream. Concerns raised included possible effects of 5FU on fertility, contraceptive requirements, and the recommendation for brief periods of abstinence following cream use to prevent partner side effects.

**Conclusions:** Although participants had some cervical cancer knowledge, misperceptions about HPV and cervical cancer were common and prevented follow-up for abnormal Pap results. Participants emphasized the need for thorough counseling about 5FU, citing this as integral for uptake. Utilizing stakeholder input to design the clinical trial is necessary to promote acceptability and adherence to the trial intervention.

## Background

Despite the availability of cost-effective prevention strategies, cervical cancer remains the second most common cancer among women worldwide.^1–3^ In sub-Saharan Africa, where the vast majority of cervical disease occurs, cervical cancer is the leading cause of cancer death among women.^3–5^ The relative scarcity of HPV vaccination and organized cervical cancer screening programs contribute to the region’s high rate of cervical cancer.^5–8^ For example, in South Africa—where our research was conducted—the age-standardized incidence rate of cervical cancer is 33 per 100,000. By contrast, the age-standardized incidence of cervical cancer in the United States and much of Western Europe is below 7 per 100,000.^9^

Sub-Saharan Africa is also the epicenter of the HIV epidemic and women living with HIV (WLWH) are far more likely to develop persistent high-risk HPV (hrHPV) infection, high-grade cervical precancer (cervical intraepithelial neoplasia grade 2 or 3 [CIN2/3)]), and invasive cervical cancer.^10–16^ Moreover, cervical precancer and cancer are harder to treat in WLWH. Standard surgical treatment for precancer (e.g., cryotherapy or loop electrosurgical excision procedure [LEEP]) is far less effective in this population.^17–20^

To meet the ambitous targets set as part of the World Health Organization’s Global Strategy to Eliminate Cervical Cancer,^21^ there is urgent need to develop novel treatment approaches for cervical precancer in WLWH. Topical therapies, such as 5-fluorouracil (5FU) and imiquimod, are currently used to treat HPV-associated precancers of the vagina, vulva, and anus. ^22–24^ There may also be a role for topical therapies—combined with surgery—in treating cervical precancer in WLWH.^25^

The overall goal of this study was to conduct formative research on the acceptability of topical 5-fluorouracil (5FU) cream as adjuvant treatment for cervical precancer in WLWH. The study was designed to inform pre-implementation planning and develop participant education materials for a planned clinical trial evaluating topical 5FU in combination with LEEP for CIN2/3 among WLWH in South Africa. Because male partner acceptance of clinical interventions, biomedical research, and contraceptive requirements has been shown to play an important role in a women’s decision to participate in research in several African settings,^26–29^ we also engaged male partners in our formative research.

## Methods

We conducted in-person, semi-structured qualitative interviews with WLWH and male partners between April 2022 and September 2022. Women were invited to participate in focus group discussions (FGDs), while male partners were in invited to participate in-depth interviews (IDIs).

FGDs and IDIs were organized at the Clinical HIV Research Unit (CHRU), a division of Wits Health Consortium and its partner NGO, Right to Care based in Johannesburg, South Africa. CHRU is co-located with Themba Lethu HIV Clinic at Helen Joseph Hospital. All interviews took place in an enclosed, confidential space and were audio-recorded, de-identified and later transcribed and translated for analysis. Participants also completed surveys providing demographic data and received reimbursement for their time and transportation to the interview site.

Research staff recruited participants from areas within and surrounding Johannesburg, including those who utilize services Themba Lethu Clinic. To be eligible for study participation, women had to be 18 years or older, living with HIV, and be willing to provide informed consent. To ensure a diverse set of experiences were represented amongst the women recruited, the study team monitored several participant characteristics during recruitment. For instance, the study team monitored the number of people being enrolled from various geographic locals to balance the number of participants coming from urban and peri-urban areas. Similarly, the team monitored the cervical cancer screening status of women to ensure adequate representation of experiences in different stages of screening and treatment. IDI recruitment occurred among male partners of WLWH; however, were not necessarily partners of the FGD participants. These men had to be 18 years or older and be willing to provide written informed consent.

For this study, definitions of acceptability and feasibility were informed by Proctor et al’s implementation science framework, which outlines outcomes for consideration when implementing interventions in new settings.^30,31^ We developed the interview guides after a review of existing literature and discussions with a multidisciplinary team, which included qualitative researchers working in South Africa. The interview guides included open-ended questions and prompts exploring participant experiences and perceptions of cervical cancer, including causes and prevention strategies, and perceptions around use of intravaginal 5FU cream following LEEP treatment for cervical precancer. We used a pelvic model demonstrate how to self-apply the 5FU cream. After several FDGs, we also solicited participant feedback on early drafts of (1) a cervical cancer informational brochure, (2) a recruitment flyer for the clinical trial, and (3) a how-to guide for intravaginal 5FU cream use.

The IDI interview guide focused on partner knowledge, attitudes, and beliefs around cervical cancer and treatment of cervical precancer. Partners were also asked about the 5FU cream and their general thoughts related to their partner’s possible participation in a clinical trial.

The interviewers were South African team members trained in qualitative data collection methods who could speak English, IsiZulu, and Sesotho, the most commonly spoken languages in Johannesburg. Informed consent was obtained prior to the interview in the participants’ preferred language. FGDs lasted approximately 90 minutes; IDIs lasted approximately 60 minutes.

Transcripts were analyzed using a rapid assessment process (RAP).^32^ We aimed to not only assess general cervical cancer knowledge, but also to inform the design and implementation of a planned clinical trial. Thus, it was imperative that we rapidly analyzed and evaluated pertinent themes and results to aid in the development of the trial. RAP allowed for a thorough qualitative analysis within a limited time frame without loss of methodological rigor.^33,34^ The analysis utilized a deductive approach in which quotations were identified and categorized into relevant domains, which had been previously utilized in designing the semi-structured interviews. These domains included knowledge, attitudes, and beliefs; acceptability; and feasibility. A RAP template was created using the domains within an excel matrix. Data analysts were trained on extracting quotations from the transcripts into the RAP template. We then pilot tested the RAP template through comparison of quotes within each domain among analysts. This allowed for consensus within each domain. Analysts were then assigned a set of transcripts from which they would extract information into the matrix by collecting quotations in a cell. Quotations were accompanied by a summary heading and short narrative description. Analysts were encouraged not to interpret but rather summarize findings. Separate matrices were used to capture data from the FDG and IDI separately. Findings from each matrix were reviewed and discussed as part of larger team meetings and key messages were formulated and extracted. Additional matrices were developed to assess how findings from women and men compared to each other.

### Ethics statement

Prior to conducting this study, we obtained ethical approval from the University of Witwatersrand Human Research Ethics Committee (M200543) and the University of North Carolina at Chapel Hill Institutional Review Board (20-1227).

## Results

We conducted 9 FGDs comprising 48 women and 18 IDIs. The mean age of participants was 43 and the majority had completed secondary education (Table 1). Among women, 75% had undergone at least one Pap smear in their lifetime and 50% had been told they had an abnormal Pap smear. The majority of these had heard of cervical cancer (80%) and 55% correctly knew the cause and how to prevent it. In contrast, only 20% of men reported they knew the cause and how to prevent cervical cancer ((Table 2). Qualitative analysis revealed three pertinent categories: factors affecting cervical cancer screening and precancer treatment, factors affecting the initiation and use of an intravaginal cream, and factors affecting adherence to an intravaginal cream.

**Table 1.** Focus group and individual interview participant demographics.

|  | Women (FGD)<br>n=48 (%) | Men (IDI)<br>n=18 (%) |
| --- | --- | --- |
| Age (mean, STD) | 43 (10) | 42 (9) |
| Highest level of education completed |  |  |
| None or some primary school | 3 (6.3) | 2 (11) |
| Completed primary school | 2 (4.1) | 0 |
| Some secondary school | 11 (23) | 8 (44) |
| Completed secondary school | 19 (40) | 6 (33) |
| Tertiary education | 13 (27) | 2 (11) |
| Relationship status |  |  |
| Never married | 26 (54) | 11 (61) |
| Married/Living together | 14 (29) | 5 (28) |
| Divorced/Separated | 3 (6.3) | 2 (11) |
| Widowed (partner has died) | 5 (10) | 0 |
| Primary medical decision maker |  |  |
| I do | 38 (79) | 8 (44) |
| I make decisions with my partner/family | 6 (13) | 8 (44) |
| My partner/family | 4 (8.3) | 2 (11) |
| A community leader | 0 | 0 |
| Primary household income source* |  |  |
| Full-time paid employment | 13 (27) | 4 (22) |
| Part-time/casual paid employment | 14 (29) | 7 (39) |
| Government grant | 15 (31) | 4 (22) |
| Financial support from outside household (e.g. other family) | 1 (2) | 2 (11) |
| No income | 9 (19) | 1 (6) |
| Personal income source** |  |  |
| Formal employment | 10 (53) | 7 (58) |
| Informal employment | 6 (32) | 3 (25) |
| Other | 3 (16) | 2 (13) |
| Type of housing |  |  |
| Brick | 39 (81) | 13 (72) |
| Informal housing (e.g. shack) | 7 (15) | 5 (28) |
| Traditional housing (e.g. hut) | 1 (2.1) | 0 |
| Other (apartment) | 1 (2.1) | 0 |
| Household water source |  |  |
| Tap water in the house | 38 (79) | 15 (83) |
| Tap water in the yard | 6 (13) | 1 (6) |
| Public tap/standpipe | 4 (8.3) | 1 (6) |
| Tap/sink at communal ablution block | 0 | 1 (6) |
| Prevalence of access to household items |  |  |
| Electricity | 47 (98) | 16 (89) |
| Toilet | 45 (94) | 16 (89) |
| Radio | 33 (68) | 9 (50) |
| TV | 42 (88) | 12 (67) |
| Mobile phone | 44 (92) | 17 (94) |
| Internet | 15 (31) | 2 (11) |

\*Participants can choose more than one option
\*\*19 women and 12 men reported earning personal income

**Table 2.** Focus group and individual interview participant self-reported experience and knowledge of cervical cancer screening.

| Questions Regarding Cervical Cancer Knowledge and Experience | Women (FGD)<br>n=48 (%) | Men (ID)<br>n=18 (%) |
| --- | --- | --- |
| Have you heard of cervical cancer before today? | 37 (77) | 10 (56) |
| If yes, how confident are you that you know how cervical cancer is caused?* |  |  |
| Not confident at all | 8 (22) | 4 (40) |
| Slightly confident | 14 (38) | 4 (40) |
| Somewhat confident | 5 (14) | 0 |
| Fairly confident | 5 (14) | 2 (20) |
| Very confident | 5 (14) | 0 |
| How confident are you that you know how cervical cancer can be prevented?* |  |  |
| Not confident at all | 10 (27) | 3 (30) |
| Slightly confident | 12 (32) | 3 (30) |
| Somewhat confident | 2 (5.4) | 0 |
| Fairly confident | 7 (19) | 2 (20) |
| Very confident | 6 (16) | 2 (20) |
| Have you ever had a screening test for cervical cancer, for example a Pap smear or an HPV test? | 41 (86) | n/a |
| If yes, has a nurse/doctor ever told you that you have abnormal cells on your Pap smear, cervical precancer, or cervical cancer? | 23 (56) | n/a |
| Have you ever received treatment for cervical precancer or cervical cancer? | 9 (19) | n/a |

\*If participants had not heard of cervical cancer before, these questions were skipped

### Knowledge, attitudes, and beliefs about cervical cancer affecting cervical cancer screening (Figure 1)

Across FGDs, women characterized cervical cancer as ‘*a cancer of the womb* [FGD #6] that needed to be addressed to avoid unhealthy outcomes. Multiple women described it as a condition which was dangerous and needed to be caught early.

> “so far what I know is that cervical cancer is a womb cancer but I do not know how dangerous it is and I don’t even know if it is curable or not that’s what I know.” (Female participant FGD #2)

**Figure 1.**
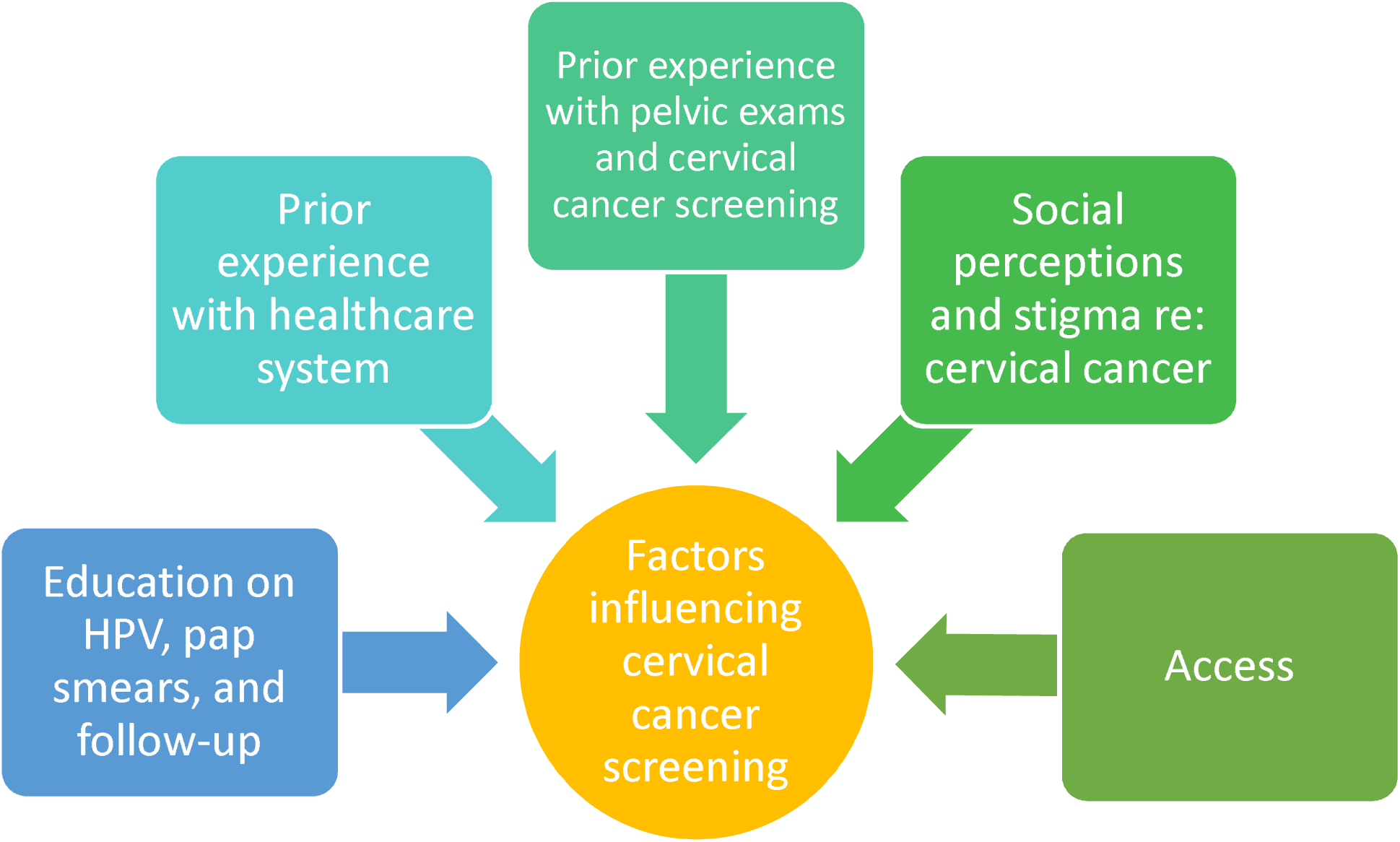
Factors affecting cervical cancer screening and follow up for abnormal pap smears.

> “Aaaah for me according to my knowledge cervical cancer is very dangerous and for us women we don’t take it serious that we have to do check up’s often. You end up discovering late, Cervical cancer is dangerous as mother [referring to another participant] has said.” (Female participant FGD #6)

By contrast, when asked about cervical cancer, many male partners were not aware of what it was even though they had heard of cancer previously. Two partners even mentioned that they weren’t aware that a cancer of the womb even existed. Male partners who were aware of cervical cancer also knew it to be a condition associated with the womb but still had questions around the causes, symptoms, and prevention.

> “Well, sometimes I would hear about breast cancer, cancer of the heart like that one for people who have heart conditions. Other cancers like the one where someone has a back problem. It depends on which cancer he/she has but this one it is the first time [I hear about cervical cancer].” (Male IDI participant)

> “I know it is cancer of the womb, you have to do Pap smear to check if everything is going well as a woman in your womb. Other than that, I do not know and I have never heard of anyone dying due to it. I heard that it is there and it is an illness that needs to be checked all the time. My partner went, I do not follow-up on ladies’ things a lot.” (Male IDI participant)

Both women and men had limited understanding of HPV and its causative role in cervical cancer. Some participants associated cervical cancer with unprotected sexual acts and sexually transmitted infections but did not name HPV specifically. The most commonly described causal pathways for cervical cancer described in multiple FGDs were the use of soaps, specific menstrual products, or the insertion of items into the vagina such as fingers, sex toys, and rags. Other causes listed by participants include genetics, lack of male circumcision, giving birth, and particular forms of contraception. When asked about cervical cancer in relation to HIV, most women and men showed a clear understanding that living with HIV increases the risk of other illnesses, including cervical cancer. When described, both groups connected cervical cancer risk to a weakened immune system caused by HIV.

> “…does giving birth to many children, does it cause cervical cancer?” (Female Participant in FGD #3)

> “And another thing that I would advise women… they want to make men happy if you go through social media these days hey things that are discussed by women no “insert snuff, do this” of which those are the things that cause cervical cancer. So they get peer pressure from people that this is how I treat my man he will never leave me.” (Female Participant in FGD #6)

> “[in response to a question on what causes cervical cancer] Aaaahh I think I don’t know if I am right, I think that illegal abortions yes….Yes, sometimes a person goes to a sangoma [traditional healer] to do an abortion so I think those can…because sometimes they are not going to do something that is done by a medical doctor I think that can also be a cause…And then some of the causes it’s not about inserting things that women put eeeehh that thing is sensitive you don’t insert the finger when you clean while bathing. It cleans itself so you will bath with soapy water you end up bathing with it at the end of the day or when years pass… those are the causes. You don’t put anything, you don’t put your hand because the hand can also give you an infection…I used to hear that eeeh like children…there are pads, we need to teach our children and ourselves not to use toilet paper.” (Female participant in FGD #6)

> “In her womb and the vagina, it [cervical cancer] might me caused by the discharge or when she is on her periods because dirty blood comes out of her.” (Male IDI participant)

> “She said it is something that starts from the womb because there is one of our cousins who had fibroids. So fibroids, I am not sure maybe they are the ones that cause cervical cancer.”( Male IDI participant)

> “Actually, I have noticed a certain pattern to it, like HIV people getting more cervical cancer but I didn’t know why exactly …that because your immune system is compromised then that cancer may develop due to that.” (Female Participant in FGD #5)

> “Knowing HIV as HIV at all. A person with HIV, their immune system is compromised that means all minor diseases can enter into her system. So cervical cancer is a big thing there is no way they cannot be vulnerable to it. So, they have…it is easy for them to get it, it is easier for them to get it.” (Male IDI Participant)

Throughout these discussions, participants also described social perceptions which influenced how they viewed cervical cancer and its causes. For instance, a WLWH felt that within the Black community there was the general perception that cancer was a ‘*white disease’* [FGD #5 2022 06 11] and that it was only just starting to impact the Black community. Expectations around sexual activity and gender intersected and influenced the way that females were expected to behave. Cervical cancer was described as a consequence when these norms were broken. For instance, when describing cervical cancer and its causes, several participants linked having multiple partners to STI and cervical cancer transmission. One male participant even described how female bodies were more adversely impacted by this than their male partners.

> “I did not hear it anywhere, it’s a thing that I am thinking that it can be this thing that can cause this cause if you compare us men and women we are not the same. It’s the same as when a woman is someone that moves [cheats] a lot, her body deteriorates quickly so a man…A person that moves a lot is a person that has different partners. So if you are a man and have different partners you won’t be somebody whose body deteriorates, you remain yourself. Not unless you have partners that are older than you, then that is where your body will deteriorate.” (Male IDI Participant)

Finally, male partners in particular, described how cervical cancer could impact their lives if their partner were to get it. These concerns predominantly focused on how cervical cancer could impact pregnancy, fertility, and their partner’s lifespan. Similar sentiments were seen among WLWH.

> “No, my concern is that maybe my partner can have it. That worries me, not knowing how long she will live. Secondly even though she had kids you will never know the future, let us say for instance we want to have a child maybe we will not be able to have. That is another concern. Then the second one is not knowing if it will go away.” (Male IDI Participant)

> “eeh womb cancer they say the womb will be having growth eeh you find that the womb has something like sores inside then it is unable to hold even if you are pregnant it is unable to hold so it may happen that you miscarry the pregnancy. You find that you miscarry and all the filth remains there, in the womb and can cause cancer in that way. You will be sick and sometimes it does not appear in your womb but your breast, breast cancer. That is what I know.” (Female Participant in FGD # 6)

> “Can cervical cancer… can I have children after I’ve had it or I’m not going to be able to have children going forward.” (Female participant in FGD #4)

When asked about how they prefer to obtain health information, women and men discussed several different modalities. These included doctors, nurses, and clinical educations. Several also mentioned traditional healers. In addition, many participants mentioned the use of brochures, pamphlets and written materials and their benefit in educating the general community. With regards to specific research study information, both men and women discussed the benefit of WhatsApp/SMS and in-person clinical education. Several male participants discussed the importance of attending appointments with their partners so they would also have an opportunity for education. When discussing the initiation of intravaginal 5FU cream, several participants thought the use a model would aid in education.

> “Brochures for example when you go to clinic they are always there, so I think they will assist a lot because people go to clinic and those process will have more information.”

> “From the clinic, we would like to hear it from clinic or hospital by a doctor, professional people.”

### Factors affecting initiation of intravaginal 5FU cream (Figure 2)

Women and male partners also shared their perceptions of research reported generally positive reviews regarding research and clinical trials. Many referred to their experiences within the context of HIV education and research. One male partner even compared the benefits of research for other health conditions to HIV research which conferred so many benefits to those living with the condition.

> “I think it is a good thing because without research you will never go anywhere. We need to actually at least try to fight this situation not every time when we see somebody with this situation you just give up….” (Male IDI participant)

**Figure 2.**
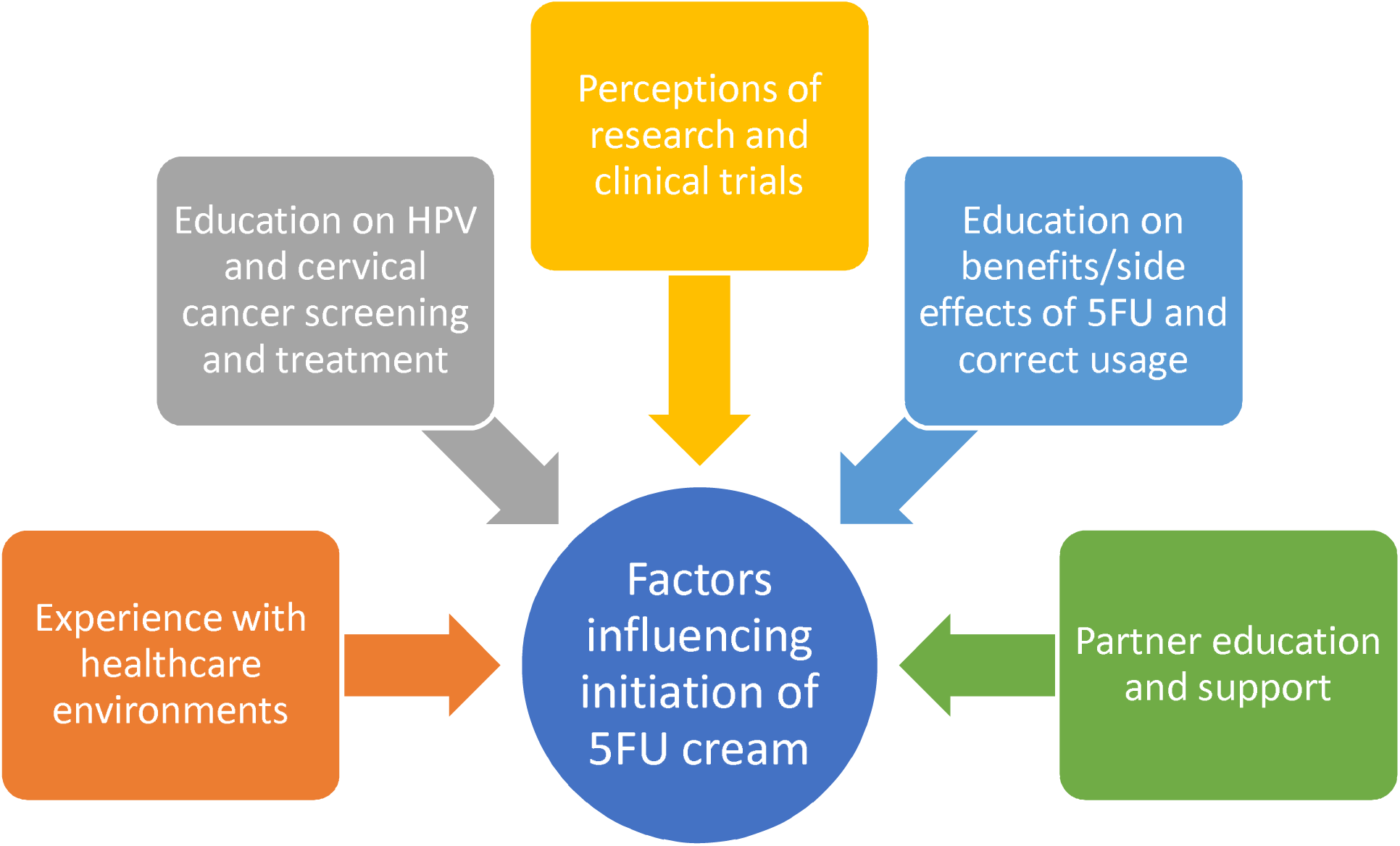
Factors affecting initiation of intravaginal 5FU cream.

> “I think that the study is the is the right way. It is good but at the same time you must know that not, it is not everyone who believes in a study. So this is like I will make an example of the vaccination. Some of us have done a vaccination, and some of us will never do it because of certain beliefs.” (Female participant in FGD #6)

In contrast, participants referenced previous negative experiences with the healthcare field outside of the research context and described their own or their partners general unwillingness to seek services at medical institutions.

> “what made me unhappy was the nurse that helped me, she was kind of moody…. She asked me where is the sticker and I told her that I forgot it and she said “did you not know that you were coming to the clinic” with attitude and I said I did know and I’m sorry. She said ok and told me to take off my panty and get on the bed but yoh the way she inserted that thing. She was not gentle so in a way it was so bad but I have let go.” (Female participant in FGD #6:)

> “Ey other are troublesome, most of us men we do not want the clinic, we do not want it. We want to go when we are really sick and we can go with an ambulance. That is when we say I can go to the hospital, but it is hard. But if you talk to the lady and have a pamphlet, she knows how to speak to him like “Baba [father] this is what we are at risk of”… (Male IDI participant)

> “No she has not been there [clinic]. She has always said she will go but tells me that she is scared.” (Male IDI participant)

Participants reported that experiencing positive interactions within healthcare was very important to them when initiating and continuing in care. Several emphasized the importance of having patient, non-judgmental, and knowledgeable staff who would maintain a high level of confidentiality and privacy. Most participants understood the importance of following medical recommendations, with many referencing their experience with HIV care.

> “It is important to know first you see, we must not judge if we can stop judging yaah they have started to know the dangers of it, the symptoms of it then you go to the clinic having those talks that are important.” (Female participant in FGD #6)

> “I am asking that they implement at the clinics and it should have privacy if I go to sister [name removed…” (Female participant in FGD #6)

> “To support her, now she did not start. She is till part of another study here and they asked if she will start taking treatment so she asked and I said treatment is a good thing. You need to take it if there is a need but I see it as a good thing and you can take it at any time” (Male IDI participant)

When asked specifically about intravaginal 5FU cream, expressed generally views after citing the potential advantages it could have. Several women saw a benefit of the cream as useful in treating pre-cancer and preventing further, more invasive, procedures, including undergoing a hysterectomy. Participants drew comparisons between how they have benefited from HIV treatment and how the cream could offer similar advantages. Male participants were generally ambivalent about cream usage and appeared to have lower comprehension about the benefits. Nonetheless, many expressed support for their partners and their participation in medical care, including use of 5FU cream.

> “I think it’s going to be very beneficial because we have been doing the LEEP thing and doing it yes they remove the cells but for me I still have the scars not inside but there are things that grow after that thing. So I was really excited when they told me about the 5FU but not for myself but for someone else but they won’t go through what I went through. Because maybe in my love life I still have to explain some things because of the things that happened which is very unfortunate because I don’t have the whole information.” (Female participant in FGD #4)

> “You know another thing that makes it easy in following you will have hope that this will cure pre-cancer. In that way you will have hope until the end because in your mind if they say you have cancer you will go crazy but when they mention the cream you would want that to remove everything. When they counsel you, they explain what the cream is and, in that way, it will be easy to do it knowing…I think you can take it like TB knowing that you can take pills and after 6 months you are done.” (Female participant in FGD #6)

While favorable views of the cream were prevalent, both men and women asked many questions about cream application and side effects. We found that concerns raised were related to application, potential impact on quality of life, and side effects. Participants additionally had questions about how to apply the cream and its purpose. Some women questioned whether it would help to alleviate existing symptoms being experienced while others wondered about the logistics of intravaginal application.

> “Yah I think trying to apply it can be a challenge, not knowing if you applied it well and it will not get off when I bath. I want to know if there won’t be an alternative like an injection?” (Female participant in FGD #6)

> “But what if someone is very sensitive of being touched by someone else, you know there are sensitive people.” (Female participant in FGD #5)

> “at first it will be a challenge, they can show you how you do it unless if they show us a structure and you look at that as to how you do it. Because some will tell you take the cream and administer it on your vagina. But how? It should be clear if we insert a finger or how do we do it when we self-administer the cream and initially it will be alright. There should be a structure that shows us how we should do it.” (Female participant in FGD #6)

### Factors affecting adherence to using intravaginal 5FU cream (Figure 3)

Facilitators provided participants with general information about how 5FU cream is used and described an 8-dose cream schedule (one dose every other week over). They also shared details of recommendations for dual contraception (condoms plus another form contraception) abstinence from sexual intercourse for 2 days following each cream application. Both women and men raised concerns regarding how the cream would impact their quality of life, particularly their sex lives. Women throughout all FGDs felt comfortable with being able to abstain for 2 days following applications and many felt their partners would be supportive, especially with appropriate clinical education. Male participants also did not raise concerns about the abstinence recommendation, although they also felt that appropriate clinical education and involvement with their partners would be integral to adherence.

> “Er what I know is that it’s very important for couples to have health talks. It will be difficult to just come out with it, but if we are used to talking it is easy to win him over. It’s easy to end up winning him over, even if he’s behind this, this, this, but if I keep showing him bit by bit, not in one day, a bit with health, and what happening outside, it’s easy for him even he doesn’t understand he will eventually be on the same page with you on issues of life, I can’t just drag him. But if you always talk about health, it’s easy to end up winning him.” (Female participant in FGD #3)

**Figure 3.**
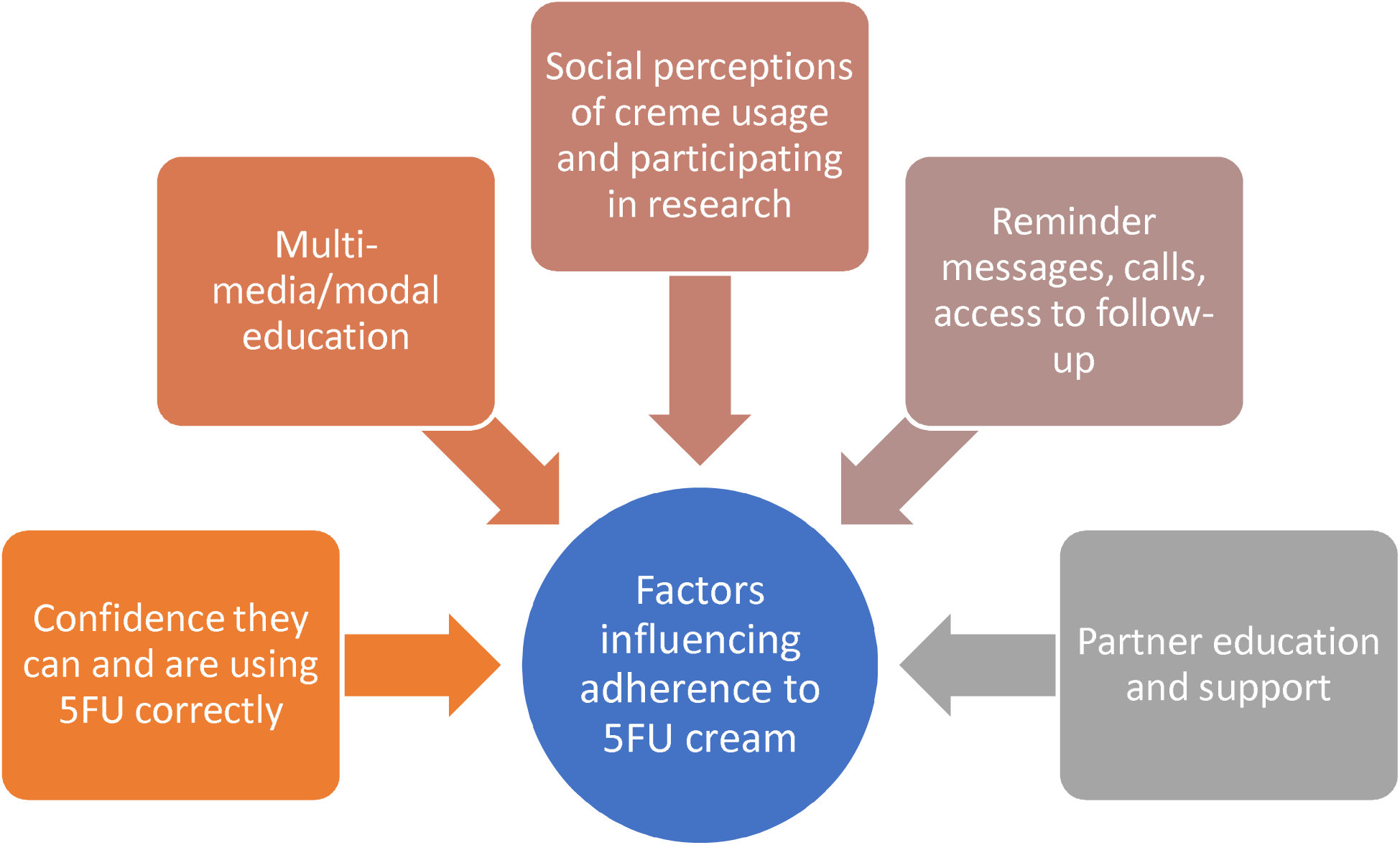
Factors affecting adherence to intravaginal 5FU cream.and support

Across several FGDs, women were concerned about not properly following guidance around cream usage and how this may impact them. One woman was concerned about being unable to adhere to the specified instructions due to issues outside of their control such as condom breakage. Another participant asked for more clarifying information around the start and end timeframe. In one FGD one participant raised the point that using a condom during the period of cream usage might be difficult for younger males due to their preferences, but they should be able to abstain from sexual activity for two days. Male partners also expressed concern over what would happen if they could not adhere the guidelines provided to them, fearing for their own health.

> “Okay so within those two days now you are applying the cream for those two days then you continue after two weeks again, do you calculate from this week Saturday Sunday then you say next week Sunday it’s two?.…Because I might mess during the two days that’s why even I have to understand if I applied for two days on Saturday and Sunday so it means that I will see it Monday, so I will not affect anyone, or the cream will not damage someone.” (Female participant in FGD #5)

> “Aaaahmm my concern is aaahm say I insert this cream then I meet with the partner we use a condom maybe let’s say the condom bursts mmmh what like…let’s say it happens at night maybe this…he will have tea…or maybe go to the clinic before 48 hours or 72 hours I want to know or maybe I have something that he can drink or just an example maybe drink a Pando it will be resolved?” (Female participant in FGD #6)

> “The cream what if a mistake happens because we do steal each other not being able to hold on anymore, so what if as a man I am unable to hold myself and we do it before 2 days ends. You see that my life is already in danger, it is scary.” (Male IDI participant)

> “…because there are these men who will say no [a surname mentioned] I have never used this thing I will not use it even today so that my concerns for women who can’t stand for themselves and can’t talk for themselves aaaahh.” (Female participant in FGD #6)

> “For the majority I think will just do the two days thing because as youngsters, the younger guys they tend to be more sexually active, so if you don’t give it to him then he goes to other people and if he goes to other people then he brings more things to kill you. The two days thing I think they’ll go for it. Waiting for two days and then be sexually active again….The condom, ehh. The majority of them don’t use condoms but if you insist maybe they will. But it’s for your own safety to use a condom afterwards till he sorts himself out and you also sort yourself out…So yes, but the majority I doubt they will use a condom.” (Female participant in FGD #5)

In addition to being concerned about how the cream would impact their quality of life, both women and male partners expressed concern over potential cream side effects. One concern raised across interviews was the potential impact of the cream and their ability to have children. A few participants were concerned that the crème would increase their chance of having children. However, the majority of participants were concerned with how the crème would impact their ability to conceive or maintain a pregnancy.

> “I’m fine right, okay now that you’ve said it, I don’t have a child at the moment. I’m more concerned what would it affect, what would be the effects on my ovaries. Yes, I’m hoping it doesn’t damage anything or cause some weird mutation that causes a deformed baby or something.” (Female participant in FGD #5)

> “Ehh, the point I’m thinking of, if it causes miscarriage or termination of pregnancy, afterwards when you’re done with the treatment [participating in the study] would your uterus or your womb still hold a child for at least full-term or just a time and then causes a miscarriage, that is the thing I’m worried about.” (Female participant in FGD #5)

> “… are they any chances that I may be fertile or can it affect anything in the cervix maybe we I have to…when I want a child and I will not get it?” (Female participant in FGD #6)

Participants were also concerned about various side effects including how the cream might affect sensitive skin, whether it would cause discharge, hair loss, itchiness, and other conditions. Throughout multiple FGDs, participants were concerned about how the cream would impact their partners, even when using condoms during sexual activity. In one FGD, a participant mentioned being concerned that her partner would leave the relationship if he got a side effect. Male partners felt similarly, expressing concern about how they would be impacted by the cream. One male partner also raised the concerned regarding if the cream would interact with HIV medication.

> “This thing of itching of the vagina will it be able to be treated is it a short term or a long-term problem that what I need to know that will it be a short term or a long term? And the other thing as they say it affects the partner does it finish his manhood or it will just be symptoms that will be shown on the partner which are those ones so that I will know that he has been affected by that cream what are the symptoms that will appear on the partner as you said it will affect the partner when we is trying to disturb me and I try to explain he says no I will not sleep so what are the symptoms for the partner.” (Female participant in FGD #6)

> “Because I might mess during the two days that’s why even I have to understand if I applied for two days on Saturday and Sunday so it means that I will see it Monday, so I will not affect anyone, or the cream will not damage someone.” (Participant in FGD #5)

> “I am concerned. My brother how can I not be concerned? But now the problem is what if… now I am thinking maybe we are having sex and the condom bursts you see, the problem that will be there. That is my concern. What if the condom bursts? So for my side how am I going to be checked? It means that I have to go to the clinic and get an injection? For STI.” (Male IDI participant)

> “Uhhm I would ask if there are no risks and whether she can continue taking her [HIV] treatment….If she is part of this study, is there any follow-up [check-up] that you will do on her to see if she won’t have problem.” (Male IDI participant)

All participants discussed the importance of community and family support in adhering to treatment. Women spoke about the support of their partners through things such as transportation and attending appointments together, emotional support, and help with reminders. Male participants desired to help with transportation to and from appointments, reminders, application of the crème itself, and emotional support. Women also discussed the importance of reminder text or instant messages. Interestingly, women tended to discuss community and family (outside of immediate partners and parents) as potentially barriers to cervical cancer screening and use of 5FU cream. This usually came up in the context of stigma and judgement around cervical cancer as potentially being a punishment.

> “I think if there’s a cream like this, it’ll help us a lot and when it comes to this part of 48 hours, I’m thinking going back to that thing of education. Education is very important not only to women, even men not even someone who is living with their boyfriend and just administered this medication…. as much as cervical cancer does not affect them directly but the other things that can affect them indirectly. Because it affects the woman, so it does affect them. And my partner has to know about it.”

> In as much as me coming as a participant, you as a nurse or counselor, you give me all the information about 5FU as to what it does. What are the side effects how do I treat the side effects, and how do I maintain and all that. Same effect as it comes with the 48 hours then I’ll get home and explain and if my partner does not understand then what do I do? Okay the first step is please give me a brochure with full information so that after laying the foundation I can give it to him. And on that brochure please make sure they are contact details so that on his free time he can manage to call you or send an email with the questions that he has I’ll probably later on make an appointment and come and see you guys maybe he’ll be comfortable to come but when I’m not there because maybe he’ll be afraid of asking the relevant questions when I’m there so rather him coming on his own.” (Female participant in FGD #4)

## Discussion

There is urgent need to improve treatment options for WLWH with cervical precancer (CIN2/3). When CIN2/3 is treated with surgery alone (as is the current standard of care), women experience high rates of disease persistence/recurrence, which in turn increases their risk of progression to cervical cancer.^17–20^ To prepare for a clinical trial evaluating topical 5FU as adjuvant treatment for CIN2/3, we sought input on the proposed intervention from WLWH and male partners.

The success of intravaginal therapies is ultimately dependent on acceptability and adherence of patients. Several failed clinical trials of vaginal microbicides underscore the central role of adherence as a core principle of successful clinical outcomes.^35–38^ For example, the CAPRISA 004 trial in South Africa reported that the impact of 1% tenofovir gel on HIV transmission was highly dependent on adherence.^39^

Using a rapid analytic approach, we assessed knowledge, attitudes and beliefs affecting cervical cancer screening and treatment of precancer in our setting. Discussions about cervical cancer and its causes across all FGDs showed that while there exists a basic knowledge of what cervical cancer is, there were still many misperceptions around its causes, prevention, and follow-up for abnormal pap smears. These are echoed in other qualitative work conducted in South Africa, which has identified a lack of appropriate counseling for abnormal pap smears and colposcopic evaluation.^40^ Our findings facilitated the development of a culturally appropriate informational brochure on cervical cancer.

FGD and IDI participants also provided insight into the salient factors likely to affect a woman’s choice to participate in our planned clinical trial and her ability to adhere to the 8-dose regimen of intravaginal 5FU cream that we aim to evaluate. Many WLWH suggested that involving their partners in the research by inviting them to attend study visits and explaining the purpose of the trial would facilitate initiation of 5FU cream and adherence to the dosing schedule.

Overall, participants were enthusiastic about topical 5FU and the proposed combination treatment approach that we plan to study. WLWH and male partners emphasized the importance of providing comprehensive patient (and partner) education and contributed to the co-creation of participant education materials for our clinical trial. Participants also felt that counselling and education would help mitigate possible barriers to sexual abstinence and condom use, as well as allay concerns regarding potential side effects.

Other potential barriers to 5FU cream use highlighted during our formative research included lack of social support, follow-up appointments requiring time away from work, cost of transportation to and from the clinic, and remembering to apply the 5FU cream at the recommended times. Although several of these potential issues are easily addresses in the clinical trial setting, they may be harder to overcome in the real world.

The strengths of this study include FDGs and IDIs that reached saturation around information. We were also able to implement participant feedback in real-time a co-create participant materials for our planned clinical trial. One of the weaknesses in this study was that the male partners were not necessarily related to the women in the FDGs thus precluding any opportunity to analyze data as dyads. Additionally, because we recruited women from health facilities, the majority had undergone cervical cancer screening at least once. Our findings may therefore not be capturing the perceptions and beliefs of women who have not previously undergone cervical cancer screening in South Africa.

In summary, acceptability of topical therapies, such as 5FU, as adjuvant treatment for cervical precancer among WLWH in South Africa is high. Barriers to initiation and adherence are multifactorial but may be mitigated through patient-centered education and, where appropriate, partner involvement. Future directions include key stakeholder IDIs to further triangulate our qualitative findings. We will also conduct a clinical trial to determine the acceptability and feasibility of topical 5FU as adjuvant treatment combined with surgical excision for CIN2/3 among WLWH in South Africa.

## Data Availability

All data produced in the present study are available upon reasonable request to the authors

## Acknowledgements

The authors acknowledge and thank Silinganiso Chatikobo for assistance during the data collection pilot and Meltonian Mzimba for assistance translating and transcribing the audio recordings from our focus groups and in-depth interviews. We are also grateful to Simone Frank and MaryBeth Grewe for their input during the design of the project.

